# The spectrum of antibacterial activity of human defensins and cathelicidin against gram-positive and gram-negative bacterial strains isolated from hospitalized patients

**DOI:** 10.1101/2020.03.03.20030890

**Authors:** Albert Bolatchiev

**Affiliations:** Department of Clinical Pharmacology, Stavropol State Medical University, Russian Federation

**Author notes:** Corresponding Author: Albert Bolatchiev, Mira Street 310, Stavropol, 355000, Russian Federation.

## Abstract

**Background:** To date, there is a spread of resistance of microorganisms to antibiotics. To solve this problem, the search and development of new drugs with antibacterial activity is necessary. Antimicrobial peptides (AMPs) have pronounced antibacterial activity and may be promising candidates for the role of new drugs. Besides, AMPs can be used to overcome conventional antibiotics resistance due to the possible synergistic effect. In this work, the combined effect of some AMPs (human defensins, HNP-1, hBD-1, hBD-3 and cathelicidin, LL-37) with conventional antibiotics (vancomycin and imipenem) against gram-positive (*Enterococcus faecalis; Staphylococcus aureus*, methicillin-sensitive, MSSA, and methicillin-resistant, MRSA) and gram-negative (*Escherichia coli*; *Klebsiella pneumoniae*; *Pseudomonas aeruginosa*) bacterial strains was investigated.

**Methods:** Bacterial strains were isolated from hospitalized patients of the intensive care unit. Commercially available AMPs (HNP-1, hBD-1, hBD-3, LL-37 by Cloud-Clone Corp., USA) and antibiotics, vancomycin (Sandoz, Slovenia) and imipenem (Merck Sharp and Dohme, USA) were used. Antibiotic resistance phenotypes of isolated bacterial strains were carried out using the disk diffusion method. The standard checkerboard assays were used to study minimum inhibitory concentrations (MIC) of antimicrobials. The combined microbicidal effect of two substances (AMP+conventional antibiotic) was assessed by the fractional inhibitory concentration index (FICI). If FICI ≤ 0,5, then it was considered that two substances showed synergism of action; if 0.5 < FICI < 4 – no interaction; if FICI > 4 – antagonism.

**Results:** All studied AMPs had antibacterial activity against the studied strains. hBD-3 showed the lowest MICs compared to other AMPs. MIC of hBD-3 against *S. aureus* (MSSA and MRSA), *E. coli, K. pneumoniae* was the same – 0.5 mg/L, and against *P. aeruginosa* it was 2 mg/L. The combinations HNP-1+vancomycin (against *E. faecalis*) and hBD-3+imipenem (against *E. coli, K. pneumoniae, P. aeruginosa*) according to FICI values have shown the synergistic effect. The results of this study can be used to develop novel antibiotics based on AMPs. Also, in some cases, AMPs can help to overcome resistance to conventional antibiotics.

## Introduction

Currently, there is a rapid and ubiquitous increase in antibiotic resistance, which is a severe problem and a challenge to modern medicine (Martinez, 2014). The threat of growing antibiotic resistance and ways to combat it are actively discussed at the level of the World Health Organization. According to WHO’s Global action plan on antimicrobial resistance, the key tasks in combating this problem are antimicrobials use optimization and new drug development (“Global Action Plan on Antimicrobial Resistance,” 2015). A decrease in the sensitivity of microorganisms to antimicrobials leads to a decrease in the effectiveness of antimicrobial therapy and, as a result, an increase in the duration of treatment, an increase in mortality and treatment financial costs (Fair & Tor, 2014; Rolain et al., 2016). So, in the USA, 19 thousand people die every year from infections caused by methicillin-resistant *Staphylococcus aureus*. Moreover, the financial costs associated with the treatment of this infection annually amount to $ 3 billion (Fischbach & Walsh, 2009). According to a recent report by the Center for Disease Control and Prevention (USA), the financial burden associated with growing microbial resistance is about $ 20 billion and 8 million extra bed days (Frieden, 2013). According to experts, by 2050, more than 10 million people will die from infections caused by resistant microbial strains every year. By this time, the world economy will lose about $ 100 trillion due to this problem (O’Neill, 2015).

The formation of antibiotic resistance is due to various reasons and mechanisms. It is known that this is a natural evolutionary process of adaptation of microorganisms to constant contact with substances with antimicrobial properties (Martinez et al., 2009). The ubiquity of antibiotic resistance is due to two factors - mutations and horizontal gene transfer (Martinez & Baquero, 2000).

The human body is continuously in contact with many non-pathogenic and pathogenic microorganisms. In the process of evolution, defense mechanisms have been formed that allow first to identify the pathogen and then, if necessary, provide adequate control of its further penetration and spread. The fulfillment of these tasks is realized through the system of innate immunity, which is able (unlike the adaptive immune system) to immediately recognize and destroy infectious agents of different natures (Iwasaki & Medzhitov, 2015). One of the essential components of innate immunity is antimicrobial peptides (AMPs) with a length of 5 to ∼100 amino acid residues. These peptides have a broad spectrum of antimicrobial activity against various infectious agents: bacteria, viruses, fungi, and protozoa. Among the six kingdoms (bacteria, archaea, protists, fungi, plants, and animals), more than 3,000 AMPs are currently identified (Wang, Li & Wang, 2016).

Among AMPs, human defensins (human neutrophil peptide-1, HNP-1; human beta-defensin-1, hBD-1; human beta-defensin-3, hBD-3) and human cathelicidin (LL-37) are of great interest, since they have a broad spectrum of antimicrobial activity (Pachón-Ibáñez et al., 2017). AMPs mechanism of action relates to its direct effect on microbial membranes, i.e., pore formation, cell leakage, and successive bacterial lysis. It is considered that the specific destruction of a microbial wall is due to the electrostatic attraction of positively charged AMPs and negatively charged membranes (Kagan et al., 1990). The spectrum of defensins and LL-37 activity is well studied (Pachón-Ibáñez et al., 2017). However, there is insufficient data on the effect of these peptides concerning bacterial strains isolated in a real clinical setting. Besides, it is crucial to study the combined antimicrobial effect of AMPs and antibacterial drugs that have long been used in the clinic.

This study aimed to determine the antibacterial activity of HNP-1, hBD-1, hBD-3, LL-37 in combination with conventional antibiotics (vancomycin and imipenem) against bacterial strains (*Enterococcus faecalis, Staphylococcus aureus, Escherichia coli, Klebsiella pneumoniae, Pseudomonas aeruginosa*) isolated from hospitalized patients.

## Materials & Methods

### Bacterial strains

*E. faecalis, S. aureus* (methicillin-sensitive, MSSA, and methicillin-resistant, MRSA), *E. coli, K. pneumoniae*, and *P. aeruginosa* strains were isolated from hospitalized patients of intensive care department of Stavropol State Regional Clinical Hospital (Russian Federation). Identification of microorganisms and their antibiotic resistance studies were carried out by EUCAST (European Committee on Antimicrobial Susceptibility Testing) protocols using the disk diffusion method (Matuschek, Brown & Kahlmeter, 2014) in The Department of Clinical Microbiology of The Center of Clinical Pharmacology and Pharmacotherapy (Stavropol, Russian Federation). According to EUCAST guidelines, the resistance of *S. aureus* to cefoxitin was considered as a marker for *mecA*-mediated methicillin resistance (Matuschek, Brown & Kahlmeter, 2014).

### Antimicrobial peptides and conventional antibiotics

Recombinant peptides: HNP-1 (purity ≥ 92%), hBD-1 (purity ≥ 95%), hBD-3 (purity ≥ 98%), LL-37 (purity ≥ 95%) (Cloud-Clone Corp., USA; peptides were expressed in *E. coli*) and conventional antibiotics: vancomycin (Sandoz, Slovenia) and imipenem (Merck Sharp and Dohme, USA) were used for checkerboard method. The amino acid sequences of the recombinant peptides are shown in *Table 1*.

**Table 1.**
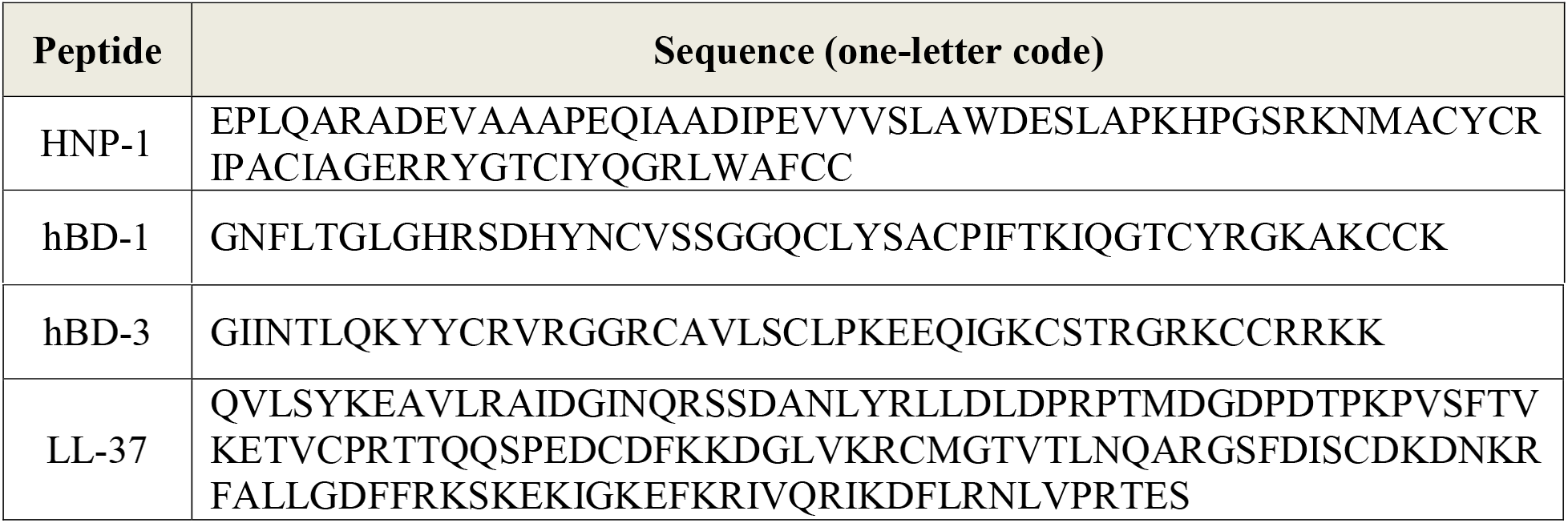
Amino acid sequences of recombinant antimicrobial peptides used in this study.

### Study of the minimum inhibitory concentrations and synergistic effects of AMPs with vancomycin/imipenem

The standard checkerboard assays were used (White et al., 1996; Orhan et al., 2005). In brief, pure bacterial cultures were cultivated on solid nutrient medium (Mannitol Salt Agar, BioMedia LLC, Russian Federation) for 18-24 hours, 37°C. From a fresh morning culture, a suspension was prepared in a sterile saline solution that corresponded to 0.5 McFarland turbidity; the resulting suspension had an approximate bacteria concentration of 1.5 *×* 10^8^ CFU/mL. Then, 0.1 mL of this suspension was dissolved in 9.9 mL of 2.1% Mueller-Hinton broth (SIFIN Institut für Immunpräparate und Nährmedien GmbH, Germany) and eventually a solution was obtained – inoculum – containing approximately 5 *×* 10^5^ CFU/mL, which corresponds to the standard EUCAST protocol (Pfaller et al., 2011).

After that, 100 µl of inoculum was administered into each well of 96-well microdilution plates (Medpolimer OJSC, Russian Federation). Then, serial twofold dilutions of each antimicrobial agent (AMP or vancomycin or imipenem, 50 µl per well) were administered into the wells. A quadruple control was performed – three wells in each microdilution plate contained: 1) only Mueller-Hinton broth; 2) only bacterial inoculum; 3) only AMPs at maximal concentrations without inoculum; 4) only vancomycin (for *E. faecalis* and *S. aureus*) or imipenem (for gram-negative bacteria) at maximal concentrations without inoculum.

For checkerboard assays, the following combinations of antimicrobials were chosen: for *E. faecalis* and *S. aureus* – AMP (HNP-1 or hBD-1 or hBD-3 or LL-37)+vancomycin; for *E. coli, K. pneumoniae*, and *P. aeruginosa* – AMP (HNP-1 or hBD-1 or hBD-3 or LL-37)+imipenem. AMPs and antibiotics were dissolved in 2.1% Mueller-Hinton broth. Vancomycin and imipenem had a dilution range from 0 mg/L to 5 mg/L. AMPs had a dilution range from 0 mg/L to 32 mg/L. The plates with the lids closed (to prevent drying) were incubated in a thermostat overnight at 37°C after adding bacterial inoculum and antimicrobials. In 20 hours, the presence or absence of growth was visually evaluated. MIC is the lowest concentration of an anti-infective agent, at which there was no visible growth of microorganisms (Milly, Toledo & Ramakrishnan, 2005).

The combined microbicidal effect of two substances (A and B) was assessed by the fractional inhibitory concentration index (FICI) (Milly, Toledo & Ramakrishnan, 2005; Ruden et al., 2009): *FICI = (A/MIC A) + (B/MIC B)*, where A and B are such concentrations of antimicrobial agents in their mixture that inhibit the growth of bacteria; MIC A and MIC B, respectively, the minimum inhibitory concentrations of substances A and B (not in combination with each other). Depending on the FICI, there are three types of mutual influence of the two investigated antimicrobials on bacteria: 1) FICI ≤ 0,5 – synergism of action; 2) 0.5 < FICI < 4 – no interaction; 3) FICI > 4 – antagonism (Odds, 2003).

## Results

Antibiotic susceptibility of isolated bacterial strains was determined using the disk diffusion method; results are presented in *Table 2*.

**Table 2.**
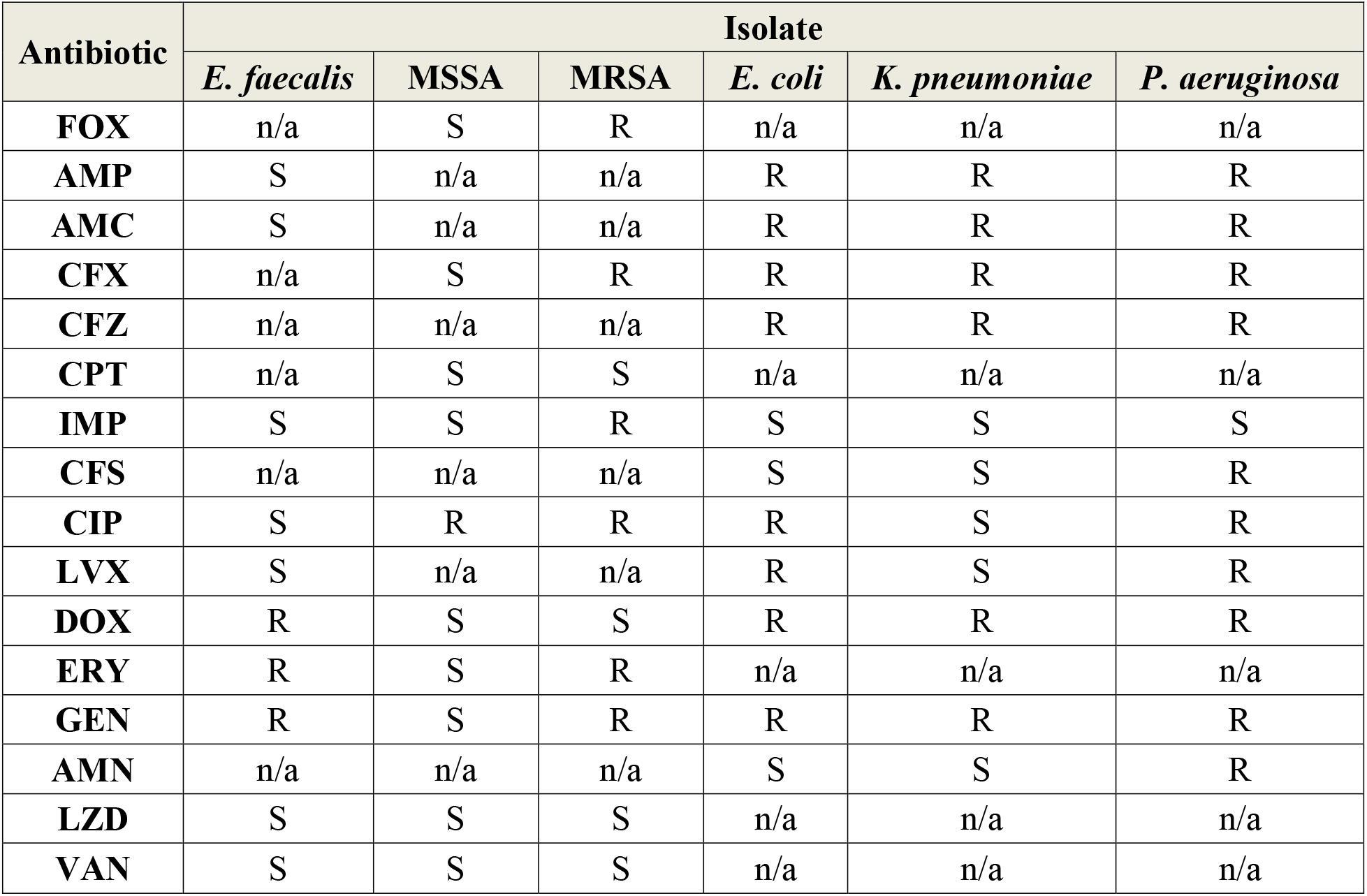
Antibiotic resistance phenotypes of isolated bacterial strains. FOX, cefoxitin; AMP, ampicillin; AMC, amoxicillin-clavulanic acid; CFX, cefotaxime; CFZ, ceftazidime; CPT, ceftaroline; IMP, imipenem; CFS, cefoperazone-sulbactam; CIP, ciprofloxacin; LVX, levofloxacin; DOX, doxycycline; ERY, erythromycin; GEN, gentamicin; AMN, amikacin; LZD, linezolid; VAN, vancomycin; R, resistant; S, susceptible; n/a, susceptibility to this antibiotic has not been determined.

The efficacy of AMPs against bacterial strains isolated from hospitalized patients of the intensive care department has been investigated *in vitro.*

*Table 3* presents the results of the MICs study. Based on the data obtained, hBD-3 had the lowest MICs against all the bacteria studied. Moreover, it is interesting that this AMP had the same MIC (0.5 mg/L) against *S. aureus* (MSSA and MRSA), *E. coli* and *K. pneumoniae*.

Combined antimicrobial effect of AMPs with conventional antibiotics has been studied using the checkerboard method (*Table 4* and *Tables S1-S24*).

**Table 3.** Minimum inhibitory concentrations of AMPs, vancomycin, and imipenem against isolated bacterial strains. VAN, vancomycin; IMP, imipenem; n/a, MIC of this antibiotic has not been determined.

| Isolate | MIC (mg/L) |  |  |  |  |  |
| --- | --- | --- | --- | --- | --- | --- |
|  | HNP-1 | hBD-1 | hBD-3 | LL-37 | VAN | IMP |
| *E. faecalis* | 1 | 8 | 1 | 16 | 0.5 | n/a |
| MSSA | 2 | 0.5 | 0.5 | >32 | 0.125 | n/a |
| MRSA | 2 | 0.5 | 0.5 | >32 | 1 | n/a |
| *E. coli* | 16 | 4 | 0.5 | 8 | n/a | 0.25 |
| *K. pneumoniae* | 16 | 8 | 0.5 | 4 | n/a | 2.5 |
| *P. aeruginosa* | 4 | >32 | 2 | 16 | n/a | 1 |

**Table 4.**
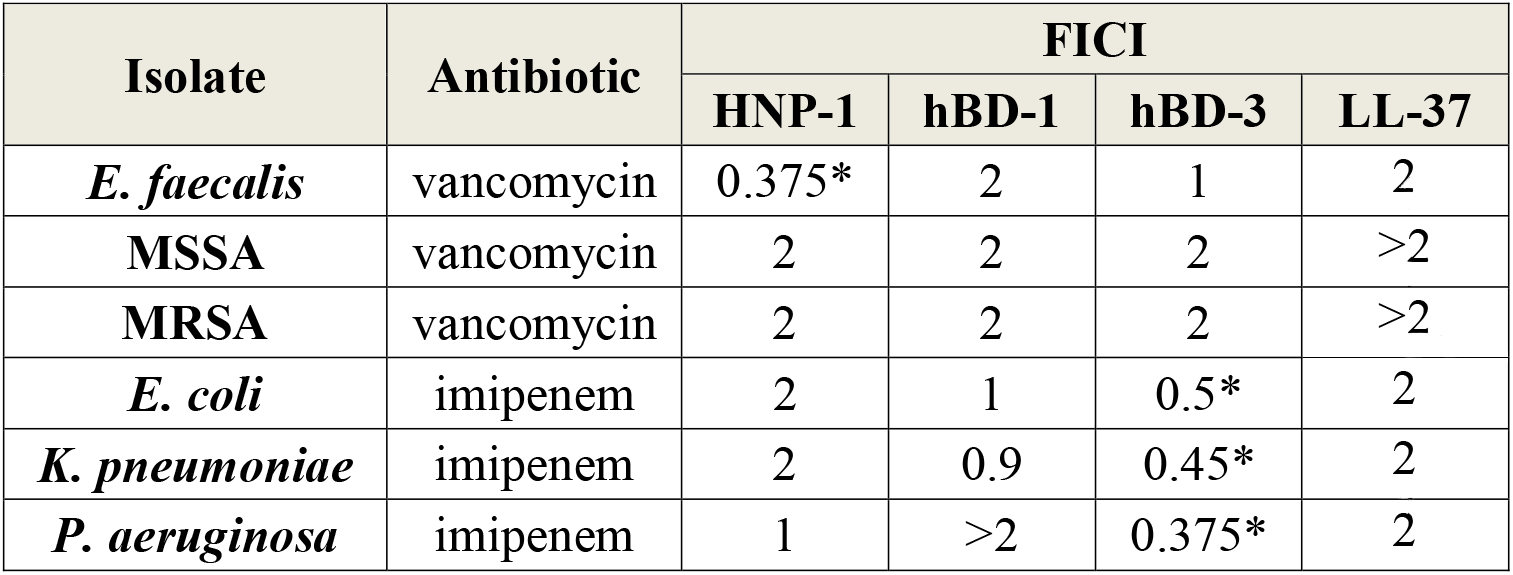
Fractional inhibitory concentration indexes (FICI) of AMPs in combination with vancomycin or imipenem against isolated bacterial strains. * – synergistic combinations.

The following combinations have shown a synergistic effect (FICI ≤ 0,5): HNP-1+vancomycin against *E. faecalis*, hBD-3+imipenem against *E. coli, K. pneumoniae*, and *P. aeruginosa*.

## Discussion

It was shown that AMPs might be effective against the studied bacterial strains *in vitro*. hBD-3 had the lowest MIC. Moreover, this peptide showed a synergistic effect in combination with the beta-lactam antibiotic imipenem against all studied gram-negative bacteria. Besides, HNP-1, in combination with vancomycin, also showed a similar outcome against *E. faecalis*. It should be noted that a lot of research is devoted to studying the spectrum of activity and the prospects for the use of AMPs. (Pachón-Ibáñez et al., 2017). The data obtained in this study are not a breakthrough in this area. However, it seemed important to investigate the effectiveness of AMPs in a clinical setting – that is, against bacterial strains isolated from severe patients being treated in the intensive care unit. Currently, defensins and LL-37 are considered promising candidates for the role of new antibiotics (Pachón-Ibáñez et al., 2017). However, the effectiveness of these AMPs varies considerably among studies and against different strains of the same species (Ganz et al., 1985; Turner et al., 1998; Schröder, 1999; Sahly et al., 2003; Dürr, Sudheendra & Ramamoorthy, 2006; Wilmes et al., 2011; Xhindoli et al., 2016). It has to be noted, that there is still no standard that defines breakpoints (criteria) of the susceptibility or resistance of specific types of bacteria to specific AMP. Although, on the other hand, perhaps this is not necessary since native AMPs are most likely not to be introduced into clinical practice. The problem of AMPs using is associated with their rapid degradation by enzymes (Lauta, 2000; Steckbeck, Deslouches & Montelaro, 2014) – this significantly affects pharmacokinetics. To solve this problem, it is necessary to modify the AMPs to increase their stability and extend the period of action.

Cases of AMPs synergy with beta-lactam antibiotics or vancomycin were shown previously (Giacometti et al., 2000; Sánchez-Gómez et al., 2011; Feng et al., 2014). Combinations of AMPs with antibiotics might be a useful tool against multidrug-resistant bacterial strains, including carbapenem-resistant gram-negative bacteria (Zharkova et al., 2019). The nature of the synergism is most likely since AMPs lead to membrane permeabilization, but conventional antibiotics usually bind to a specific target in the bacterial cell (Zharkova et al., 2019). Thus, AMPs and antibiotics have a different mechanism of action, but this does not always lead to a potentiation of the effect (Bolatchiev et al., 2020). Perhaps this is because AMPs have an alternative, unknown mechanism of action. However, it should be noted that *in vivo* AMPs exhibit different biological effects. AMPs directly stimulate the migration of immune cells, promote the release of pro-inflammatory cytokines and activate antigen-presenting cells via Th1-immune response – thus, AMPs are an effective link between innate and adaptive immunity (Suarez-Carmona et al., 2015). In this regard, it is crucial to study the effectiveness of AMPs *in vivo* since they not only have a direct antimicrobial effect but also stimulate the immune response.

## Conclusions

All studied antimicrobial peptides (HNP-1, hBD-1, hBD-3, LL-37) have shown antibacterial activity against *E. faecalis, S. aureus* (MSSA and MRSA), *E. coli, K. pneumoniae*, and *P. aeruginosa* strains isolated from patients of intensive care unit. hBD-3 had the lowest values of the minimum inhibitory concentration in comparison with other studied peptides. Also, hBD-3 demonstrated a synergistic effect in combination with a beta-lactam antibiotic imipenem against gram-negative strains. The combination of HNP-1 with vancomycin showed a similar effect against enterococci. The results obtained can be used to develop new antibiotics based on antimicrobial peptides. Moreover, the data collected may allow developing strategies to overcome resistance to conventional antibiotics through the additional use of antimicrobial peptides. Further, *in vivo* studies are needed to confirm this hypothesis.

## Data Availability

All of the data referred to in the manuscript is available

## Funding

**This work was funded by Russian Foundation for Basic Research according to the research project No. 18-315-00081**

## Acknowledgements

Many thanks to Professor Vladimir Baturin for constructive comments about the manuscript. Thanks to Dr. Elena Kunitsina for providing bacterial strains. Big thanks to the directorship of Stavropol State Medical University represented by the rector Professor Vladimir Koshel and the vice-rector Professor Evgeny Shchetinin for providing excellent facilities for research.

## Notes

### Competing Interest Statement

The authors have declared no competing interest.

